# Switched forced SEIRDV compartmental models to monitor COVID-19 spread and immunization in Italy

**DOI:** 10.1101/2021.06.21.21259230

**Authors:** Erminia Antonelli, Elena Loli Piccolomini, Fabiana Zama

## Abstract

This paper presents a new hybrid compartmental model for studying the COVID-19 epidemic evolution in Italy since the beginning of the vaccination campaign started on 2020/12/27 and shows forecasts of the epidemic evolution in Italy. The proposed compartmental model subdivides the population into six compartments and extends the SEIRD model proposed in [E.L.Piccolomini and F.Zama, PLOS ONE, 15(8):1–17, 08 2020] by adding the Vaccinated population and framing the global model as a hybrid-switched dynamical system. Aiming to represent the quantities that characterize the epidemic behaviour from an accurate fit to the observed data, we partition the observation time interval into sub-intervals. The model parameters change according to a switching rule depending on the data behaviour and the infection rate continuity condition. In particular, we study the representation of the infection rate both as linear and exponential piecewise continuous functions. We choose the length of sub-intervals balancing the data fit with the model complexity through the Bayesian Information Criterion. The calibration of the model shows an excellent representation of the epidemic behaviour and thirty days forecasts have proven to reproduce the infection spread reliably. Finally, we discuss different possible forecast scenarios obtained by simulating an increased vaccination rate.

## 1. Introduction

Compartmental models are essential mathematical tools in the analysis of the evolution of epidemics, for prediction and simulation of future strategies which can be used by governments and policymakers to allocate sanitary and economic resources. The parameters of such models are related to meaningful characteristics of the epidemic disease, such as infection rate, infectious period, lethality rate. Moreover, through such models, it is possible to estimate the number of secondary cases produced by a single infected person at start time (basic Reproduction number *R*_0_) and during the epidemic evolution (effective time-dependent Reproduction number *R*_*t*_). In particular, the trend of *R*_*t*_ is of great importance to check the epidemic evolution over time.

The COVID-19 pandemic, caused by the Sars-CoV-2 virus, has renewed interest in studying these models and a significant number of papers appeared on this subject since the beginning of 2020 (refer to LitCovid database for up to date literature [1]). They differ each other for the type of model proposed, the external events considered, such as movement restrictions imposed by governments or quarantines, and the regions where models are applied.

Starting from the first SIR (Susceptible (S), Infected (I), and Recovered (R)) model, proposed in 1927 by Kermack and McKendrick [2], several generalizations have been formulated over the years by increasing the number of compartments, such as, for example, the Susceptible – Exposed – Infectious – Recovered (SEIR) and the Susceptible - Exposed - Infected - Recovered - Dead (SEIRD) schemes. Further extensions have been proposed to model the COVID-19 outbreak considering the different social distancing policies and control measures applied in the various geographic areas to contain the epidemic spread. More compartments have been added, making the models more and more complex (see [3], [4], [5], [6], to mention only a few of the most recent).

In this paper we intend to consider the effects of the vaccine on the epidemic spread by further extending the SEIRD model adding the compartment of Vaccinated people, thus obtaining a SEIRDV scheme. Among the vaccine-related papers within the COVID-19 literature several hypothetical scenarios are analysed based on different prioritisation policies according to vaccine efficacy and its availability [7]. Other papers focus on the possible benefits of combining vaccination with Nonpharmaceutical Interventions (NPIs) such as surveillance, social distancing, social relaxation, quarantining, patient treatment/isolation (see [8, 9] and references therein).

Following the approach in [10], we introduce a switching rule that governs the SEIRDV model state at any given time. Besides producing optimal fit to epidemic data, introducing such a hybrid approach allows us to represent disease evolution when restriction policies and virus variants cause changes in fundamental parameters such as infection rate, recovery periods, and death rates. Although switched models are widespread in various engineering applications, studies about epidemic models are less common; see, for example, [11] (SIRV), [12](SIR) and [13] (SEIRD). In particular the authors in [13] propose a hybrid SEIRD model with a mortality rate represented by an inverse exponential function where the residual correction is based on the ARIMA method. The model, tested on US COVID statistic data in the period February-September 2020, made precise predictions for up to 2 months ahead. The reader can also refer to [13] for an exhaustive bibliography.

Concerning the model parameters, it is well known that COVID-19 epidemic data cannot be accurately represented by any compartmental approach with constant parameters all over the epidemic duration. To face this problem, some authors use variable parameters in the time interval (for example [6]) or change the fitting function (see [5]). In our approach all the model parameters are constant in each switching time interval, except for the infection rate which is a time-dependent forcing function modelled as piecewise continuous.

The model calibration is carried out by solving a sequence of constrained minimisations of the weighted least-squares residuals between the measured epidemic data and the value of the state variables, which satisfy the initial value ordinary differential system representing the SEIRDV model.

This paper is an extension of our previous work [14], where we proposed a SEIRD model (before the availability of vaccines), with two different forcing functions, to monitor the first phase of the evolution of Covid-19 in Italy (2020/02/24-2020/05/24). Compared to [14] we modify the model, by including the Vaccinated compartment (we remark that the vaccination campaign started in Italy on 27 December 2020), by representing the proposed model into the hybrid models’ theoretical frame, and by changing the expression of the forcing functions. Moreover in the calibration phase, we add weights in the fitting objective function and bound constraints, thus improving the model computational effectiveness and accuracy.

In the experimental section, we report the results obtained by our scheme on the Italian national and regional epidemic data in the period 2020/12/27-2021/06/12. The inclusion of two different expressions for the infection rate function allows us to obtain different possible scenarios which prove to be very useful in the prediction phase.

### 1.1. Contributions

We summarize here the main contributions of this paper.

- We propose a SEIRDV scheme by adding the Vaccinated compartment to the well known SEIRD model.
- We consider a dynamical switched framework where the interval length is chosen on the basis of the Bayesian Information Criterium.
- We represent the infection rate as a continuous time dependent function comparing a linear and an exponential piecewise formulation.

The rest of the paper is organized as follows. In Section 2, we present the proposed model and the calibration procedure. The Section 3 reports the calibration results for the data of the COVID-19 in Italy and in the Emilia-Romagna region as well as possible forecast scenarios. Conclusions are drawn in Section 4.

## 2. METHODS

This section introduces the details of the proposed switched compartmental model and its algorithmic formulation. We start by describing the SEIRDV model in paragraph 2.1, then we introduce the switched model together with insights about the time-dependent infection rate functions (paragraph 2.2). In paragraph 2.3, we discuss the algorithmic details of the calibration procedure consisting of a sequence of bound-constrained optimization problems defined by the model switches. Finally, in paragraph 2.4 we briefly discuss how we use the model to make predictions.

### 2.1. The SEIRDV model with constant parameters

The SEIRDV model characterized by constant parameters is obtained from the SEIRD model [14] by adding the new compartment *V* representing the vaccinated population. The following differential system represents the populations’ dynamics:

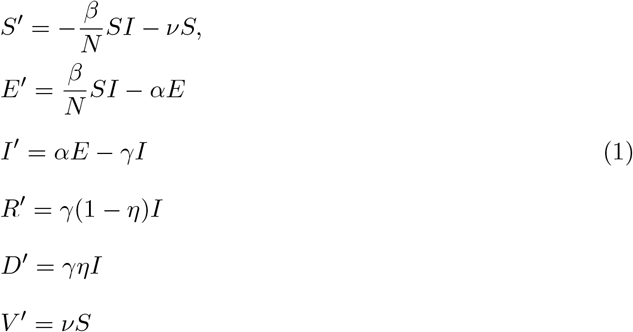

where the total population, assumed of constant size *N*, is subdivided into six compartments: Susceptible (S), Exposed (E), Infected (I), Recovered (R), Dead (D) and Vaccinated (V). The system (1) is solved starting from an initial time *t* = *t*_0_ where the values *S*(*t*_0_), *E*(*t*_0_), *I*(*t*_0_), *R*(*t*_0_), *D*(*t*_0_), *V* (*t*_0_) are assigned on the basis of the available data and integrated up to a final time *T*.

The parameter *β* ≥ 0 represents the infection rate, accounting for the susceptible people infected by infectious people. Its value is related to the number of contacts between Susceptible and Infected. Standard models, as well as our SEIRDV, assume this relationship to be linear. The parameter *α* > 0 represents the incubation rate for the transition from Exposed to Infected states. Such value relates to the incubation period *A*_*I*_ as follows: *A*_*I*_ = 1*/α*. The average incubation ranges from 2 to 14 days (*d*) (see https://www.worldometers.info/coronavirus/coronavirus-incubation-period/). According to [15], more than 97 percent of people who contract SARS-CoV-2 show symptoms within 11.5 days of exposure. Recently a comparative study assesses the incubation period around 6.5 days [16]. The cited studies assess the value of *α* in the interval [0.14, 0.5].

The parameter *γ* > 0 representing the removal rate relates to the average infectious period *T*_*I*_ as *γ* = 1*/T*_*I*_. At the beginning of the outbreak, an average value *T*_*I*_ ≃ 20*d* has been measured [17], hence *γ* ∈ [0.03, 0.1].

After the period *T*_*I*_, the Infected split into Recovered and Dead with weights 1 − *η* and *η* respectively (0 ≤ *η* ≤ 1). Hence the parameter *η* represents the fraction of the Removed individuals who die and its value depends on environmental situations that change over time, such as the population age, the virus spread, medical care availability and treatments. Finally, the parameter *ν* > 0 represents the vaccination rate. Its value is particularly useful in the prediction phase to obtain different scenarios.

Important information about the epidemic development is obtained from the number of infection cases generated from a single infectious individual, i.e. the basic reproduction number *R*_0_, defined as follows (see details in appendix Appendix B):

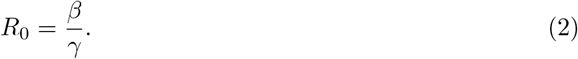

It is well known that the epidemic occurs when *R*_0_ > 1; however, this information refers to the initial stage, assuming that the entire population is Susceptible. In the case of COVID-19, estimations of *R*_0_ in the interval [1.5, 6.68] were obtained during the first months of 2020 [18].

### 2.2. Switched forced SEIRDV model

The movement restriction policies adopted worldwide as well as the occurrence of different virus variants cause changes in the value of the infection rate *β* and possibly of other model parameters over time. In order to monitor the model parameters from the measured data flexibly, we propose a *hybrid switched* version of the SEIRDV model and represent the infection rate *β* as a continuous time-dependent function, modelled according to the epidemic data. We split the time interval [*t*_0_, *T*] into *p* sub-intervals Δ_*k*_ = [*t*_*k*−1_, *t*_*k*_] (*k* = 1, …, *p* and *t*_*p*_ = *T*) and define a switching rule Θ setting the values of the model parameters as follows:

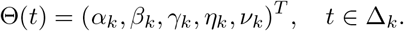

Then the hybrid model is represented as [10]:

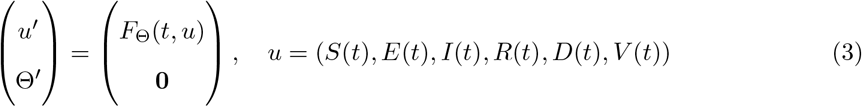

with state variable (*u*, Θ)^*T*^ and *F*_Θ_(*t, u*) as in (1), and with model parameters represented by the piecewise constant function Θ(*t*).

However, using a constant value for the infection rate does not represent the epidemic behaviour in a sufficiently flexible way [14]; therefore, we introduce a continuous time-dependent infection rate ***β***(*t*). In this case, the epidemic model is known in the literature as *forced* model (see for example [19] chp 6). In this paper, we represent the infection rate as piecewise linear and exponential interpolating functions, yielding to SEIRDV_pwl and SEIRDV_pwe models, respectively.

Let us define ***β***_*k*_ (*t*) the restriction of ***β***(*t*) to the interval Δ_*k*_, *k* = 1, …, *p*, and set the values *β*_*k*_ ≡ ***β***(*t*_*k*_), *k* = 0, …, *p*. The SEIRDV pwl defines the infection rate as:

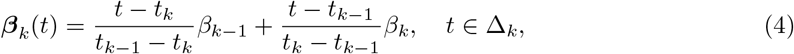

whereas SEIRDV_pwe defines the infection rate as follows:

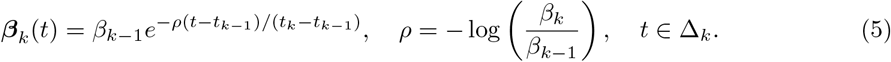

We observe that for both models it holds :

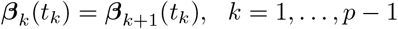

hence ***β*** is continuous in [*t*_0_, *T*].

The evolution of the global hybrid forced model, represented in figure 1, shows the changes of the epidemic model at each switching interval Δ_*k*_ represented by the values of the model parameters defined as Θ_*k*_ ≡ Θ(*t*), *t* ∈ Δ_*k*_. The restriction of the dynamical model (3) on each interval Δ_*k*_, is represented in Figure 2, where the model populations, for *t* ∈ Δ_*k*_, are given by (*S*_*k*_, *E*_*k*_, *I*_*k*_, *R*_*k*_, *D*_*k*_, *V*_*k*_).

**Figure 1:**
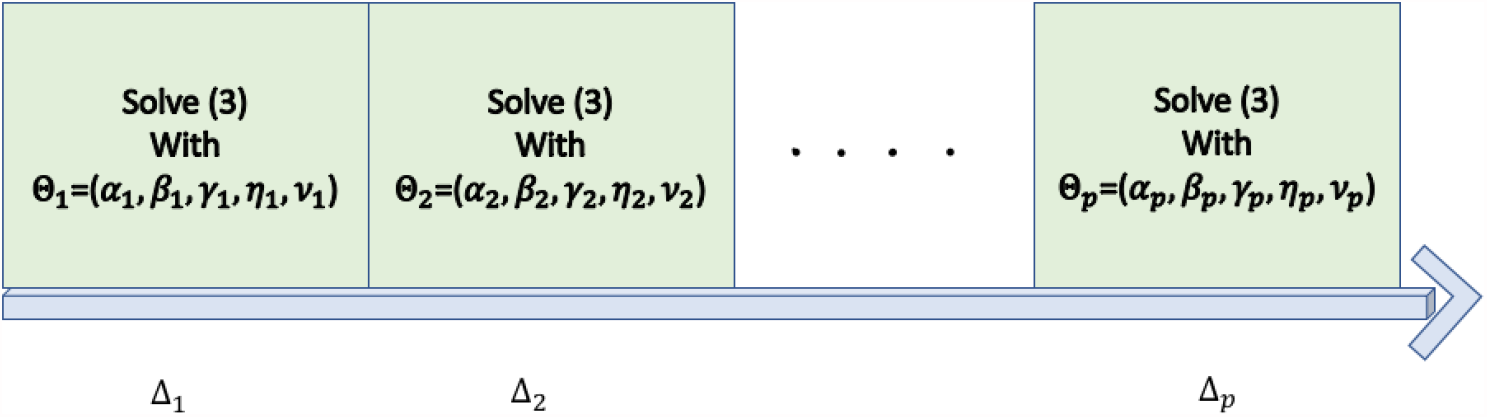
The evolution of the global hybrid model, related to the values of the parameters Θ_*k*_.

**Figure 2:**
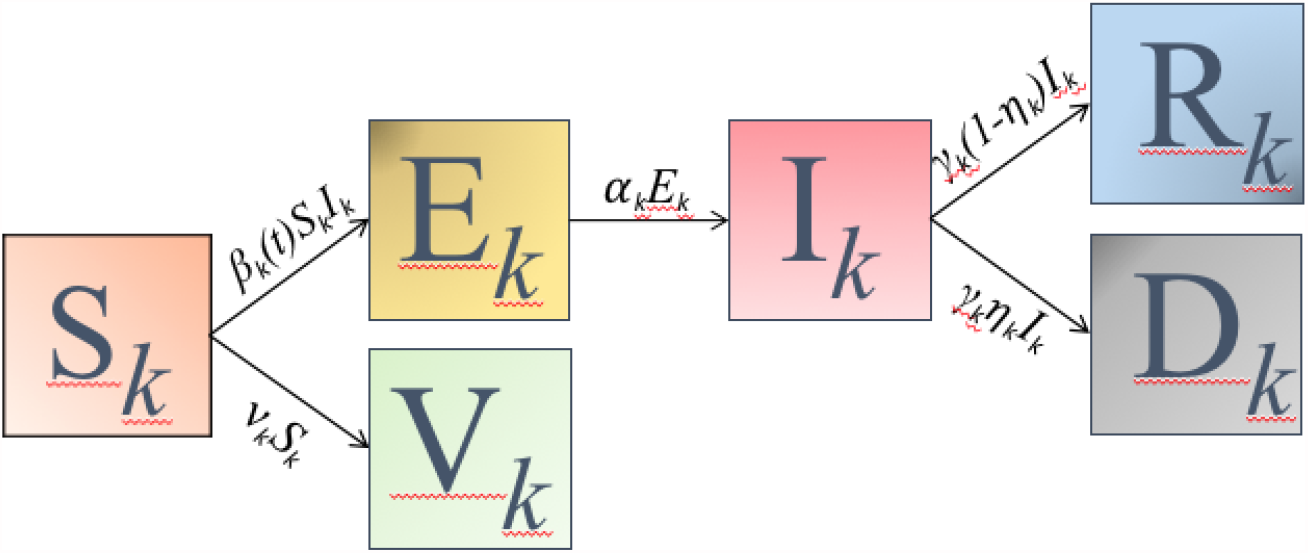
Dynamical model (3) restricted to the interval Δ_*k*_.

### 2.3. Calibration Procedure

This paragraph describes the calibration procedure in the interval [*t*_0_, *T*] supposed of *n* days. We calibrate the parameters on the sub-intervals Δ_*k*_, *k* = 1, … *p* with uniform size *L* = ⌊*n/p*⌋ (special care is taken in the case *n ≠ L · p* to avoid a too small length of Δ_*p*_).

Generally, we are interested in keeping *L* as large as possible to guarantee a proper balancing between the data fit and the model complexity, evaluated in terms of the number of parameters to be calibrated. In section 3 we discuss the details of the choice of a proper value for *L*.

We describe now the parameter estimation process in a single sub-interval Δ_*k*_. We collect the observed data about Infected, Recovered, Dead and Vaccinated compartments in vectors I,R,D and V of size *L* and stack them into the matrix *Y* ∈ℝ^*L×*4^, *Y* = [*I, R, D, V*]. Then we consider *Z* ∈ℝ^*L×*4^ as the restriction of *u*(*t*; Θ) (defined in (3)) to the components *I*(*t*), *R*(*t*), *D*(*t*), *V* (*t*) computed in the measurement days in Δ_*k*_ and we compute the model parameters Θ_*k*_ solving a weighted constrained nonlinear least squares problem of the form:

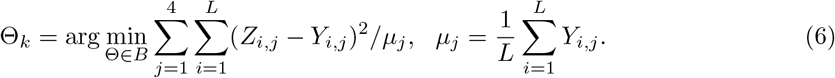

where the positive weights *µ*_*j*_ are introduced to compensate different data scales. The bound set *B* is defined as

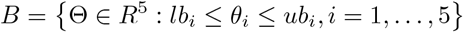

where the upper bounds *ub*_*i*_ = 1, *i* = 1 …, 5 and lower bounds

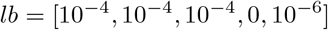

contain the values estimated in the literature.

To solve the minimization problem (6) numerically, we use iterative solvers as discussed in section 3. Figure 3 schematically represents the calibration steps of the global hybrid model in the whole interval [*t*_0_, *T*]. The scheme highlights that the results Θ_*k*_ of the minimization problem on Δ_*k*_ is taken in input as starting guess in the minimization problem on Δ_*k*+1_.

**Figure 3:**
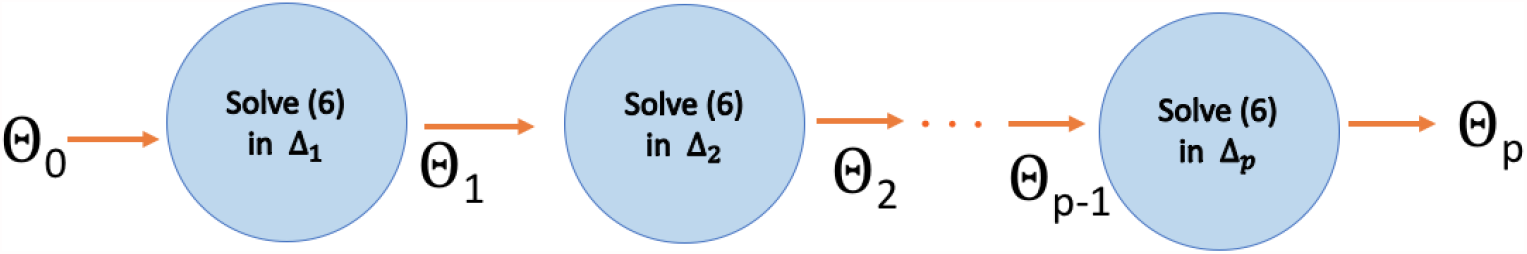
Calibration steps of the global hybrid model parameters Θ_*k*_.

To suitably choose the first starting guess Θ_0_, which has a fundamental role in the quality of the final solution, we solve problem (6) on a unique short time interval *T*_*i*_ of about ten days with starting guess (0.1, 0.1, 0.1, 0.1, 0.1).

In Algorithm 1 we summarize the main steps of the calibration phase to estimate the parameters Θ_*k*_, *k* = 1 …, *p*.

#### Algorithm 1

Calibration Algorithm

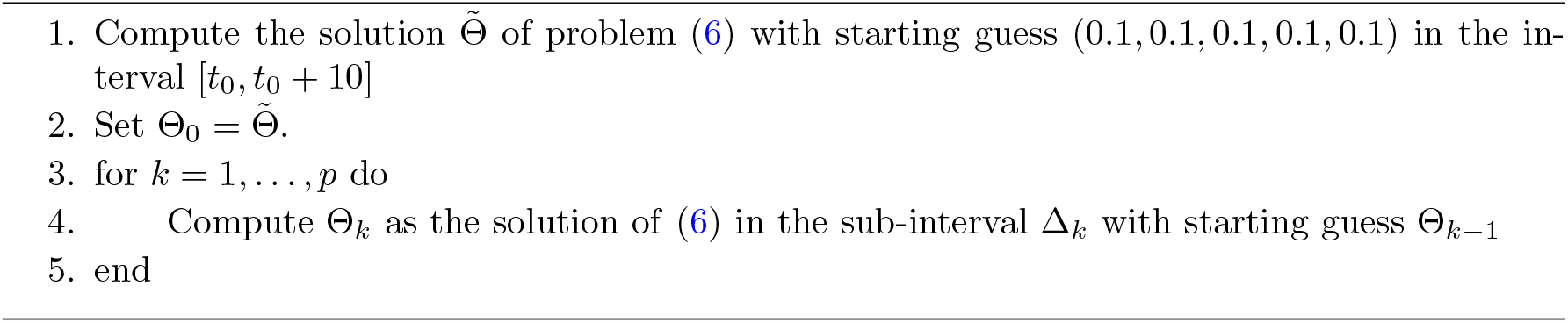

The forward differential problem (1) is solved by a fourth order variable step Runge-Kutta method. The initial conditions in each sub-interval Δ_*k*_ are given by the observed values of the compartments *I, R, D, V* in the initial day of Δ_*k*_. Concerning the starting value *E*_0_ of the Exposed compartment, which is not available from data, in the starting interval Δ_1_, we relate it with the delay time *t*_*d*_ between the contact with the infectious agent and the onset of symptoms or signs of infection, as follows:

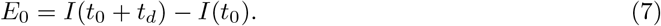

(see Section 3 for more details on the values of *t*_*d*_). In the successive intervals Δ_*k*_, *k* > 1, we set *E*_0_ as the last value of Exposed computed in the interval Δ_*k*−1_. The starting value of Susceptible is the difference between the total population *N* and the sum of all the other compartments.

### 2.4. Prediction

We use SEIRDV to predict the future behaviour of the disease evolution in short-medium *m*-days interval [*T, T*_*m*_], with *T*_*m*_ = *T* + *m*. In this paper we have adopted the following two strategies for prediction.

1. We set in (1) the parameters Θ_*p*_ = (*α*_*p*_, *β*_*p*_, *γ*_*p*_, *η*_*p*_, *ν*_*p*_) computed in the last calibration interval Δ_*p*_ and we run the model over a unique time interval [*T, T*_*m*_]. In our simulations we use both the SEIRDV_pwl and SEIRDV_pwe proposed approaches.
2. We set in (1) the parameters Θ_*σ*_ = (*α*_*p*_, *β*_*p*_, *γ*_*p*_, *η*_*p*_, *σ · ν*_*p*_), with *σ* > 1, to simulate an increased vaccination rate. We compute also in this case the prediction using both the linear and exponential *β* functions.

## 3. Numerical results

The results presented in this section have been obtained by implementing the SEIRDV_pwl and SEIRDV_pwe algorithms in Matlab 2021a. The codes are available on https://github.com/fzama63/COVID-SEIRDV.

### 3.1. Data description

Epidemic data are downloaded from the repository open source Github https://github.com/pcm-dpc/COVID-19 of the Italian Civil Protection Department, containing the official data provided by the Ministry of Health (see [20] for a detailed description). We consider here the global national data (*N* = 60360000) as well as the regional data from Emilia-Romagna (*N* = 4445900).

Information about vaccine administration is obtained in the Github repository: https://github.com/italia/covid19-opendata-vaccini (see details in Appendix A).

### 3.2. Model calibration

We have solved the constrained least squares problems (6) by means of the `lsqnonlin` Matlab function from the `Optimization Toolbox` with the trust-region reflective algorithm. The initial value differential problem (1) has been solved with the `ode45` Matlab function.

To analyse the numerical solution of the calibration in the interval [*t*_0_, *T*] constituted of *n* days, we compute, for any considered population, a Relative RESidual defined as:

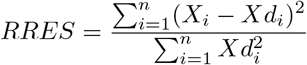

and the Bayesian Information Criterion (BIC) [21], defined as follows:

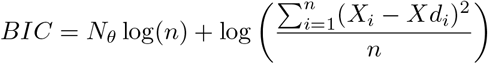

where *N*_*θ*_ is the number of the estimated parameters, *Xd*_*i*_ represents the acquired compartment data and *X*_*i*_ is the corresponding value computed by the calibrated model at day *i, i* = 1, …, *n*. The *BIC* takes into account the number of model estimated parameters and tends to penalize the inclusion of additional parameters. The lower this quantity, the better the model will be.

To set a convenient initial value *E*_0_ for the Exposed compartment we run SEIRDV_pwl and SEIRDV_pwe on the first 15 days, from 2020/12/27 to 2021/01/10, choosing *E*_0_ as in (7) with *t*_*d*_ = 1, …, 10 days. Then we compute *RRES* for the Infected compartment and choose *t*_*d*_ corresponding to the minimum value. As shown in figure 4, the smallest *RRES* is obtained when *t*_*d*_ = 2 for both SEIRDV_pwl and SEIRDV_pwe, hence we continue with *t*_*d*_ = 2 throughout this section.

**Figure 4:**
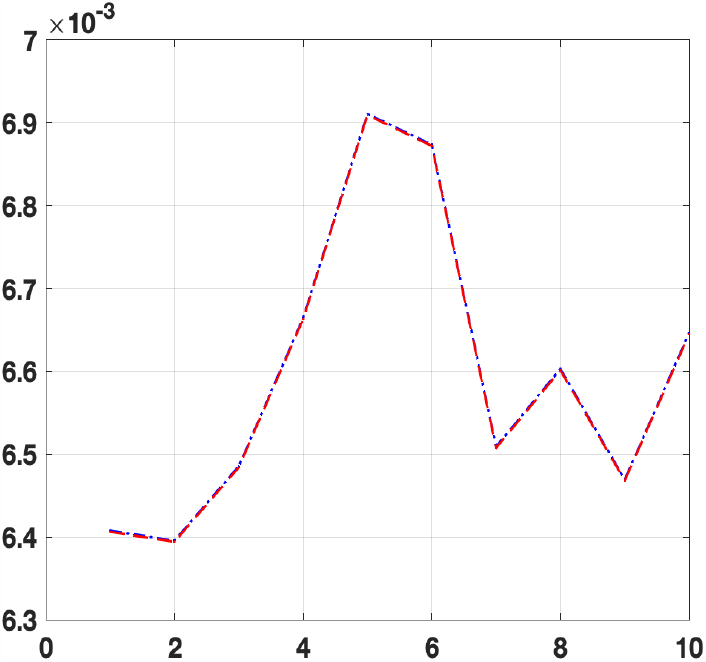
*RRES* vs *t*_*d*_ for SEIRDV_pwl (blue dash-dotted line) and SEIRDV_pwe (red dashed line)

We define the calibration period of *n* = 85 days, from 2020/12/27 to 2021/3/21 (we remind that in Italy vaccination campaign started on 2020/12/27) and run the calibration of SEIRDV_pwe and SEIRDV_pwl models.

The first aspect of our analysis concerns the choice of the number of switches. We split the whole time interval into sub-intervals of fixed length *L* (except for the last one which can be of different size). By increasing their number, we decrease the fit error at the expense of the computation cost, determined by the number of parameters, proportional to the number of switches. In order to choose the best value *L* we try all the values in the interval [5, 85] days, we compute the minimum *BIC*_*min*_ over all BIC values and we evaluate: Δ_*BIC*_ = *BIC*_*X*_ − *BIC*_*min*_ where X is any of the considered populations (I,R,D or V).

In Figure 5 we plot the value Δ_*BIC*_ vs *L* for each compartment: red dot-dashed line for SEIRDV_pwe and blue line for SEIRDV_pwl model. Except for the Vaccinated (figure 5(d)), all the populations show the minimum *BIC* for *L* = 21.

**Figure 5:**
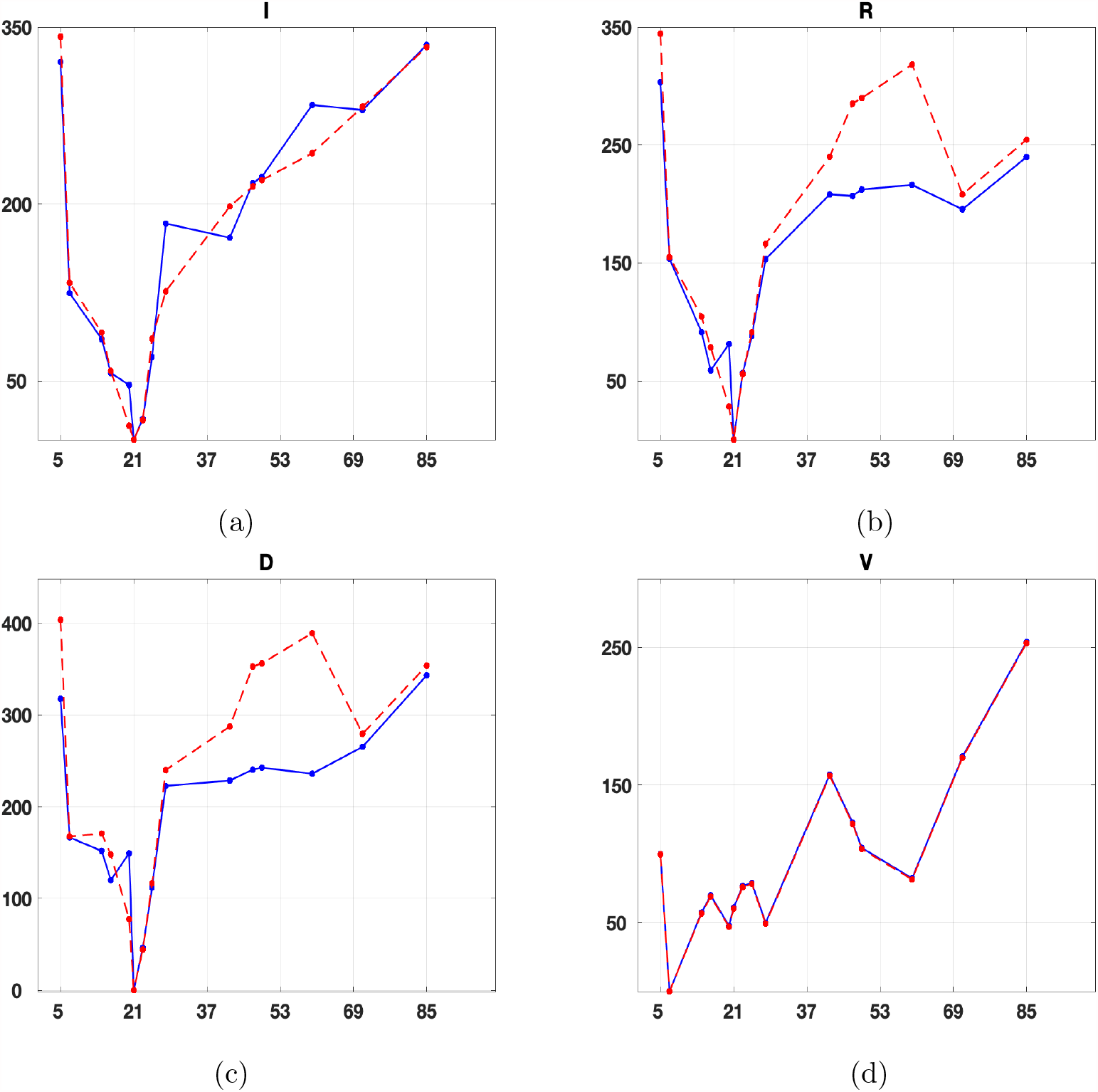
Δ_*BIC*_ vs *L* for each compartment: red dot-dashed line for SEIRDV_pwe and blue line for SEIRDV_pwl model. (a) Infects (b) Recovered (c) Dead (d) Vaccinated.

Hence throughout this section, we split the calibration time into four sub-intervals of length *L* = 21 obtaining the following time intervals:

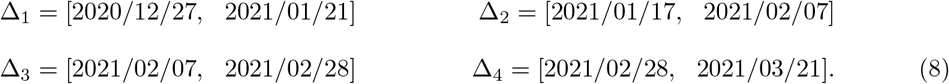

We calibrate the model parameter using Algorithm 1. For the solution of the constrained optimization problem, we compared the `trust-region reflective` (TR) method with the Levemberg Marquardt (LM) one, which overperforms the Broyden-Fletcher-Goldfarb-Shanno (BFGS) as noted in [5]. We report in Table 1 the number of function evaluations FCount and the relative residual *RRES* for the Infected, Recovered, Dead and Vaccinated compartments. We find that for both exponential and linear infection rates, the TR method is computationally the most efficient (smallest number of function evaluations) and it is also slightly more precise than LM. Therefore we continue our analysis applying the TR method.

**Table 1:**
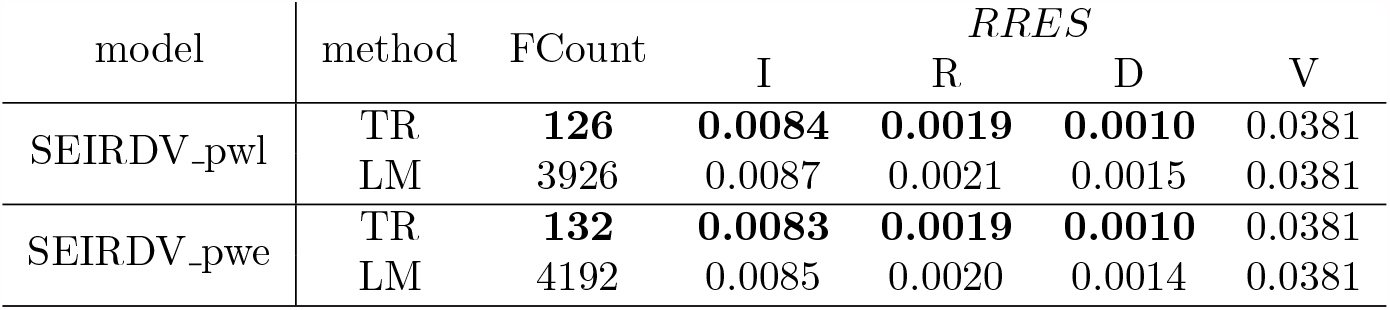
Number of function evaluations FCount and relative residual *RRES* obtained by LM and TR solvers for SEIRDV_pwl and SEIRDV_pwe. In bold the best results obtained for each algorithm.

Figures 6 (a) and 7 (a) and (b) plot the calibrated functions of Infected, Recovered and Vaccinated populations, respectively. We can appreciate the good quality of data-fit of SEIRDV_pwl and SEIRDV_pwe. In Figure 6 (b) we represent the difference between the infected population obtained by SEIRDV_exp and SEIRDV_pwl algorithms. We observe that the main differences occur in the Δ_3_ and Δ_4_ intervals.

**Figure 6:**
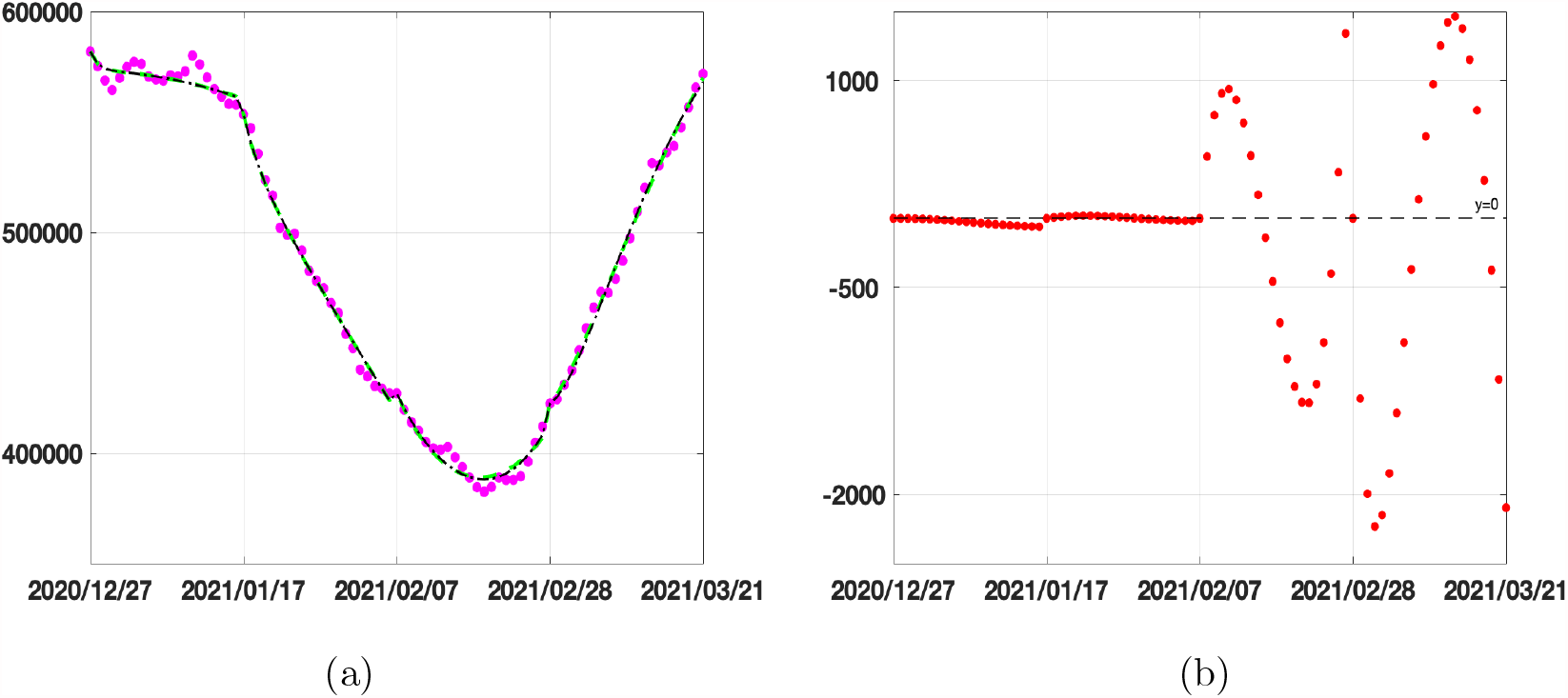
Calibration results on Infected compartment. (a) Data from 2020/12/27 until 2021/03/21 (magenta circles), SEIRDV_pwl calibration (black dashed line), SEIRDV_pwe calibration (green continuous line). (b) Difference between SEIRDV_pwe and SEIRDV_pwl (red circles).

**Figure 7:**
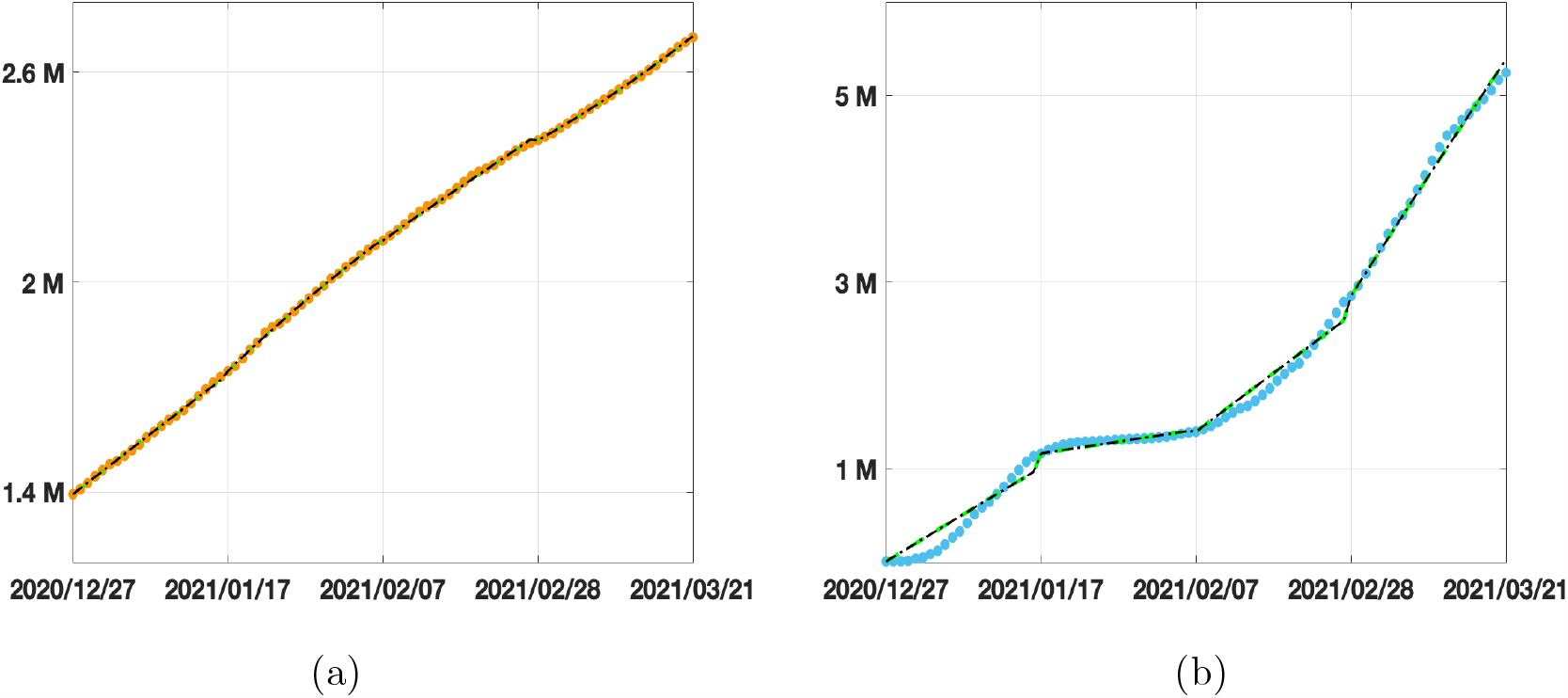
Calibration results. (a) Recovered compartment. Data from 2020/12/27 until 2021/03/21 (orange circles), SEIRDV_pwl calibration (black dashed line), SEIRDV_pwe calibration (green continuous line). (b) Vaccinated compartment. Data from 2020/12/27 until 2021/03/21 (cyan circles), SEIRDV_pwl calibration (black dashed line), SEIRDV_pwe calibration (green continuous line).

In Figure 8 we plot the values of both the infection rate function *β*(*t*) (on the left) and the reproduction number function *R*_*t*_(*t*) (on the right) computed as follows

**Figure 8:**
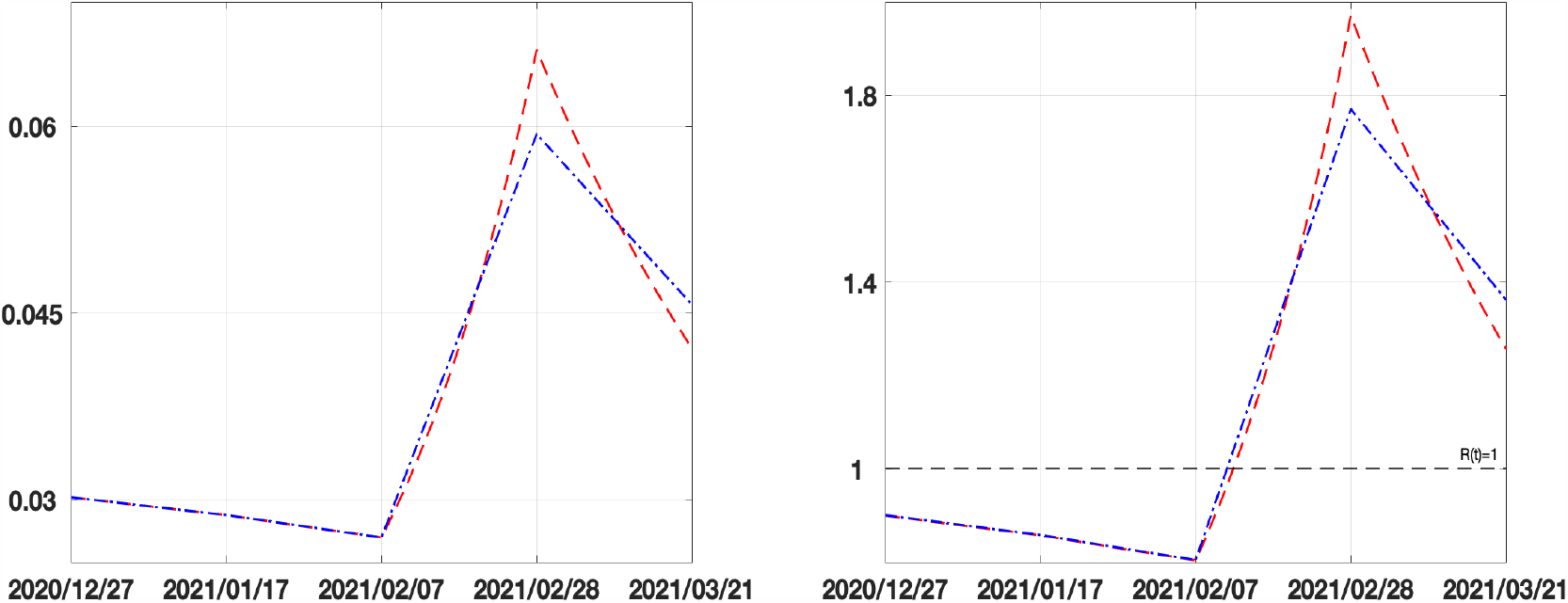
Forcing *β* functions (on the left) and Reproduction index *R*_*t*_ (on the right) for SEIRDV_pwl (blue dash-dotted line) and SEIRDV_exp (red dashed line) models.

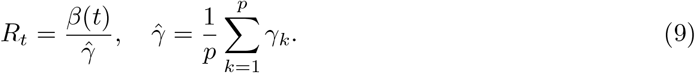

The red line relative to the exponential model changes more rapidly when the epidemic spread increases (Δ_3_ and Δ_4_ intervals). To analyse the behaviour of the infection rate in the considered sub-intervals, we average the values of the calibrated function *β*(*t*), represented in figure 8 (a), on the intervals [Δ_1_, Δ_2_], getting 0.00286 for both the methods, and in the period [Δ_3_, Δ_4_] obtaining 0.0480 and 0.0487 for SEIRDV_pwl SEIRDV_pwe, respectively. These values show that both methods capture the increase of the infection rate that causes the inversion of the epidemic curve in [Δ_3_, Δ_4_].

We now discuss the parameters calibrated by Algorithm 1 and reported in Table 2. Concerning the incubation rate *α*, both models report a decreasing behaviour in the period Δ_1_ − Δ_4_. It corresponds to an incubation period between 2 and 4 days. The removal rate *γ* is very similar for both methods and gives the following removal periods: 34 *d*(Δ_1_), 25.9 *d*(Δ_2_), 26.9 *d*(Δ_3_), 34.2 *d*(Δ_4_) producing the average removal period of 30.3 *d*.

**Table 2:** Parameters calibrated in the different time intervals (8).

| parameter | model | $\Delta_1$ | $\Delta_2$ | $\Delta_3$ | $\Delta_4$ | mean |
| --- | --- | --- | --- | --- | --- | --- |
| $\alpha$ | SEIRDV_pwl | 0.9790 | 0.4293 | 0.3217 | 0.2571 | 0.4968 |
|  | SEIRDV_pwe | 0.9790 | 0.4297 | 0.3433 | 0.2276 | 0.4949 |
| $\gamma$ | SEIRDV_pwl | 0.0294 | 0.0386 | 0.0371 | 0.0293 | 0.0336 |
|  | SEIRDV_pwe | 0.0294 | 0.0386 | 0.0372 | 0.0293 | 0.0336 |
| $\eta$ | SEIRDV_pwl | 0.0290 | 0.0238 | 0.0216 | 0.0238 | 0.0245 |
|  | SEIRDV_pwe | 0.0294 | 0.0386 | 0.0372 | 0.0293 | 0.0336 |
| $\nu$ | SEIRDV_pwl | 0.0008 | 0.0002 | 0.0010 | 0.0022 | 0.0011 |
|  | SEIRDV_pwe | 0.0008 | 0.0002 | 0.0010 | 0.0022 | 0.0011 |

Regarding the parameter *η*, we observe that the average value 2.4% obtained by SEIRDV_pwl slightly underestimates the reference value 3%, reported by Johns Hopkins University Coronavirus Resource Center, https://coronavirus.jhu.edu/data/mortality. On the contrary, the average value 3.2% obtained by SEIRDV_pwe constitutes a slight overestimate. Both models compute the same vaccination rate *ν* in each period.

### 3.3. Prediction on national data

In this paragraph, we apply the calibrated SEIRDV_pwl and SEIRDV_pwe to make predictions. To test the forecast reliability, we compute a prediction in the interval Δ_5_ = [2021/03/21, 2021/04/20] using the data available in that period to measure the precision of our forecast in terms of the Infected peak time and value.

In Figure 9 we show the predicted Infected curve. With the red dashed curve we plot the prediction obtained by using the values of all the parameters calibrated in the last interval Δ_4_. With the continuous blue line, we plot the prediction obtained by changing only the *β*(*t*) function as the curve interpolating the infection rate calibrated in 2021/02/28 and 2021/03/21 (drawn with a blue and a red star, respectively). Comparing the prediction curves relative to SEIRD_pwl (Figure 9 (a)) and SEIRD_pwe (Figure 9 (b)) with the epidemic data represented by magenta empty circles we can see that the exponential model is more accurate.

**Figure 9:**
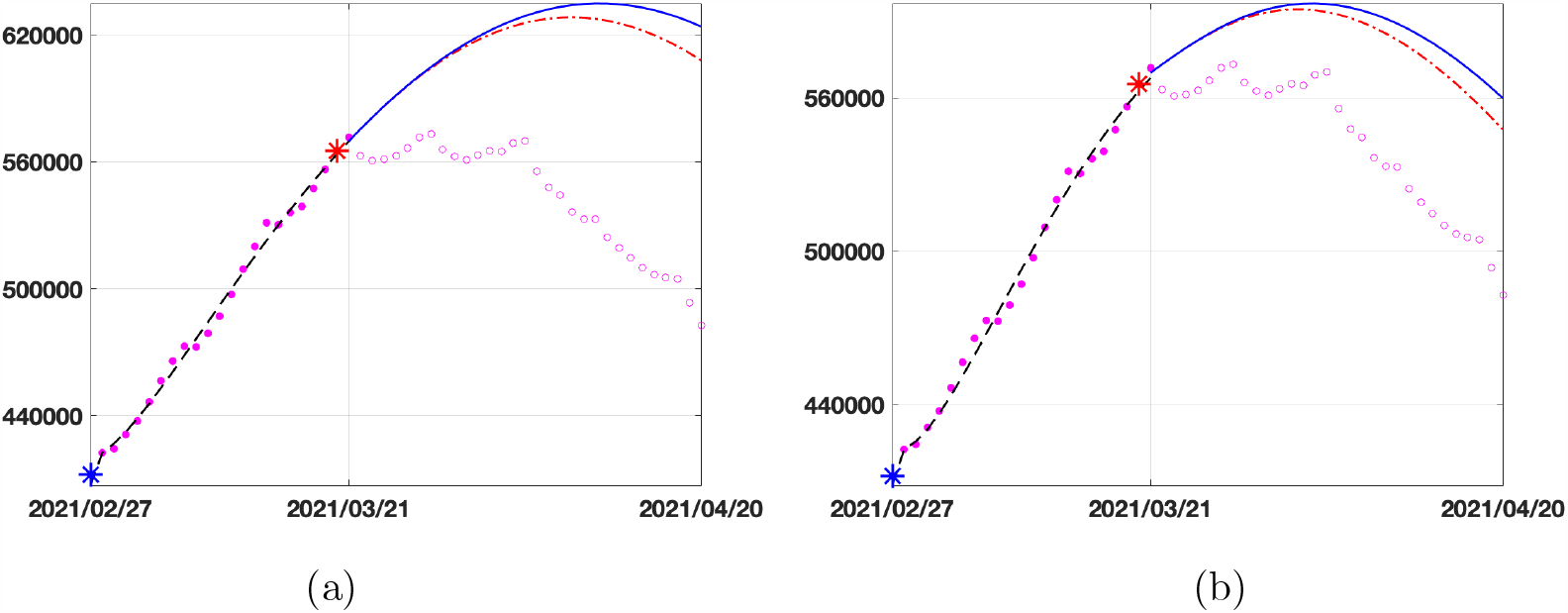
Prediction for the Infected compartment in Italy from 2021/03/22 to 2021/04/20. Data (magenta circles), prediction with *β*(*t*) calibrated in the last interval Δ_4_ (red dashed line) and prediction with *β*(*t*) interpolating the the values of *β* in red and blue stars (blue continuous line). (a) SEIRDV_pwl (b) SEIRDV_pwe

We highlight that the reported forecast refers to the vaccination rate *ν* = 0.0022, computed in the calibration interval Δ_4_ corresponding to 133572 administration per day. From Table 3 we can see that the peak of infected people is reached on 2021/04/09 and 2021/04/03 with SEIRD_pwl and SEIRD_pwe predictions, respectively. We observe that SEIRD_pwe curve is closer to the available data that reaches its maximum on (2021/03/28) with 573235 Infected people.

**Table 3:** Results of the prediction experiment in Italy obtained with different vaccination rates: number of Infected people and day of the peak for SEIRDV_pwl and SEIRDV_pwe. The peak of available data is on 2021/03/28 with 573235 Infected people.

| vaccination<br>rate | administration<br>per day | SEIRDV_pwl |  | SEIRDV_pwe |  |
| --- | --- | --- | --- | --- | --- |
|  |  | #(Infected) | peak day | #(Infected) | peak day |
| 0.0022 | 133572 | 628504 | 09/04/21 | 594604 | 03/04/21 |
| 0.0055 | 333930 | 622139 | 07/04/21 | 592549 | 01/04/21 |
| 0.0088 | 534288 | 617322 | 05/04/21 | 590882 | 01/04/21 |

In the second and third lines of the table, we report the maximum number of Infected and the day of forecast peak increasing the vaccination rate. To graphically represent the effects of an increased vaccination rate, we plot in Figure 10 the results given by the two models in 40 days using the vaccination rates in Table 3. Comparing the two models, we see that the SEIRDV_pwe gives the more realistic prediction.

**Figure 10:**
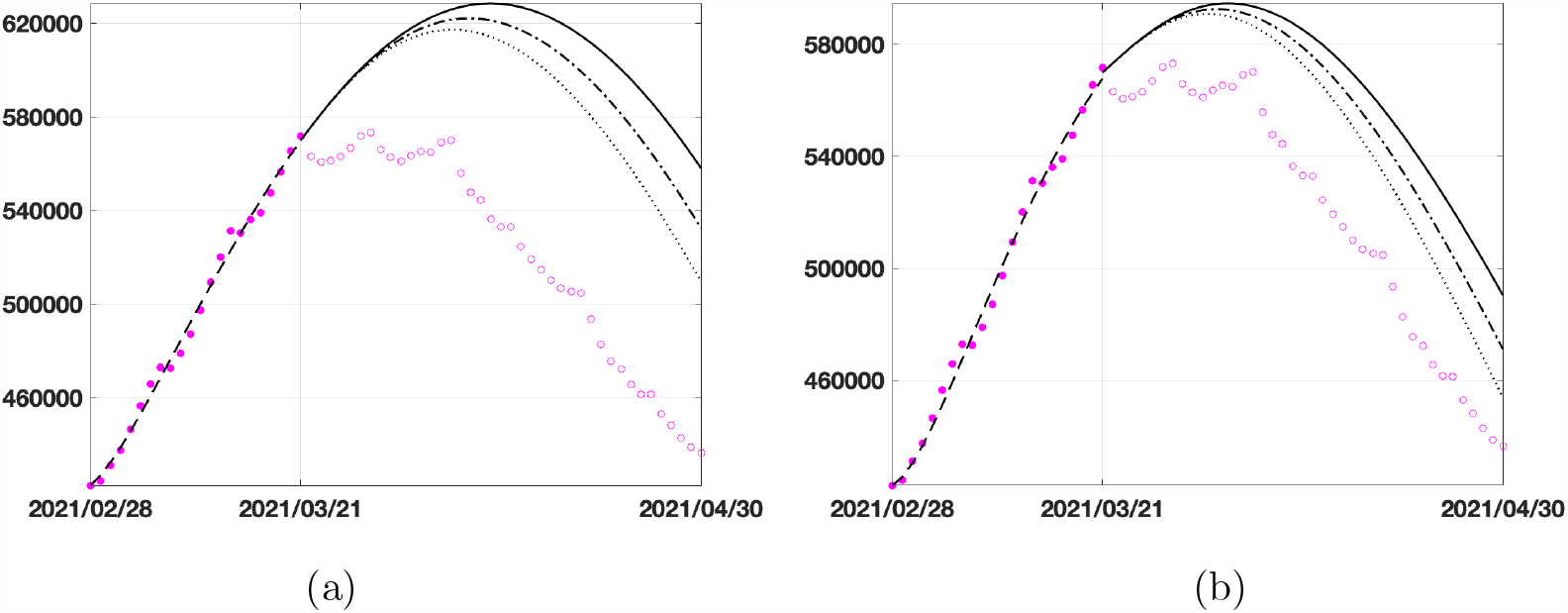
30 days prediction for the Infected compartment in Italy by considering different vaccination rates *v* = 0.0022129 (dark continuous line), *v* = 0.0055323 (dark dashed-dotted line) and *v* = 0.0088517 (dark dotted line) : (a) SEIRDV_ pwl, (b) SEIRDV_pwe

### 3.4. Prediction on regional data

Finally, we present the prediction obtained using more homogeneous and smaller-scale data acquired in the Emilia-Romagna region after performing the calibration on the same sub-intervals as in (8). In Figure 11 we plot the prediction obtained with the same procedure as in Figure 9. Differently to what happens for the Italian case, in the linear model (Figure 11 (a)) the prediction obtained with the red curve is entirely inaccurate, whereas for the exponential model (Figure 11 (b)) the red and blue lines define a region containing the Infected data. Therefore, SEIRDV_pwe can be used to make reliable predictions with both strategies. Finally, in Figure 12 we show the results for increasing vaccination rates, as done in Figure 10 for the national data. The SEIRDV_pwl forecasts are now closer to the Infected data compared to SEIRDV_pwe, confirming the importance of having both methods available to make different predictions.

**Figure 11:**
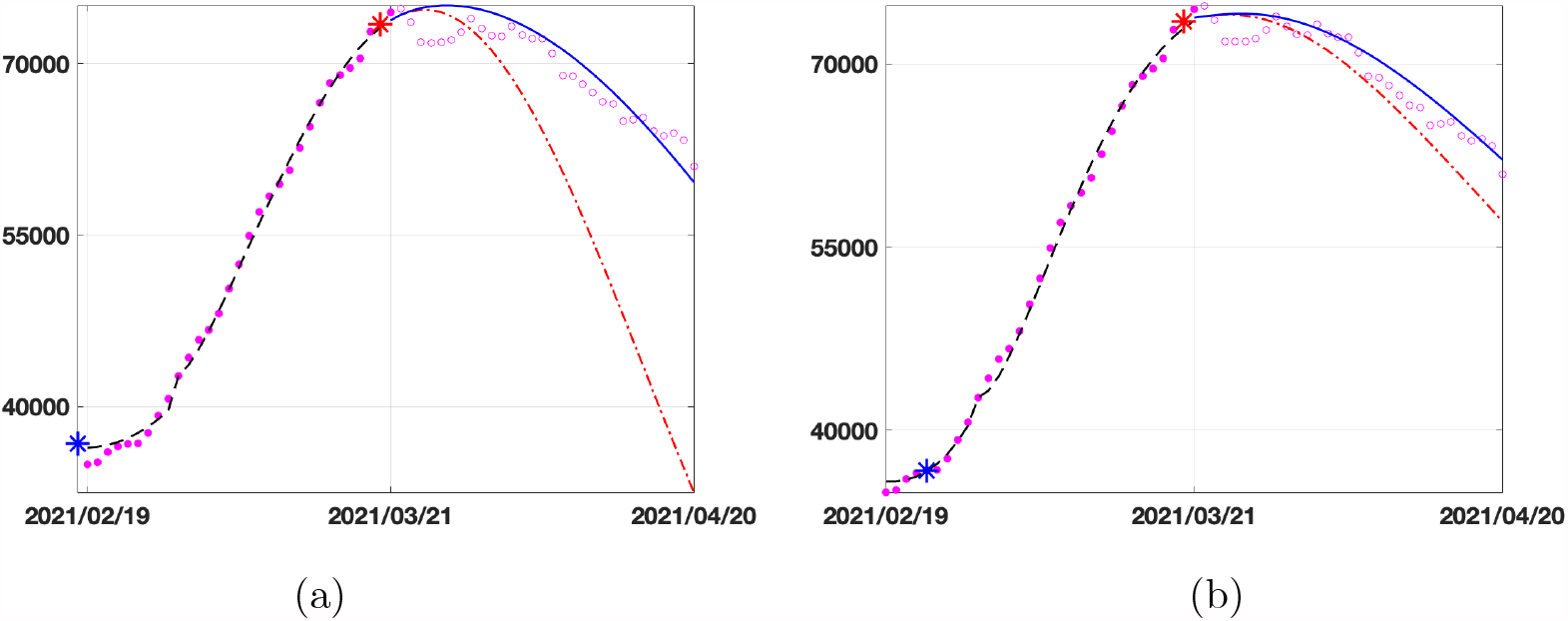
Prediction for the Infected compartment in Emilia-Romagna from 2021/03/22 to 2021/04/20. Data (magenta circles), prediction with *β*(*t*) interpolating the the values of *β* in red and blue stars (blue continuous line), prediction with *β*(*t*) calibrated in the last interval Δ_4_. (a) SEIRDV_pwl (b) SEIRDV_pwe

**Figure 12:**
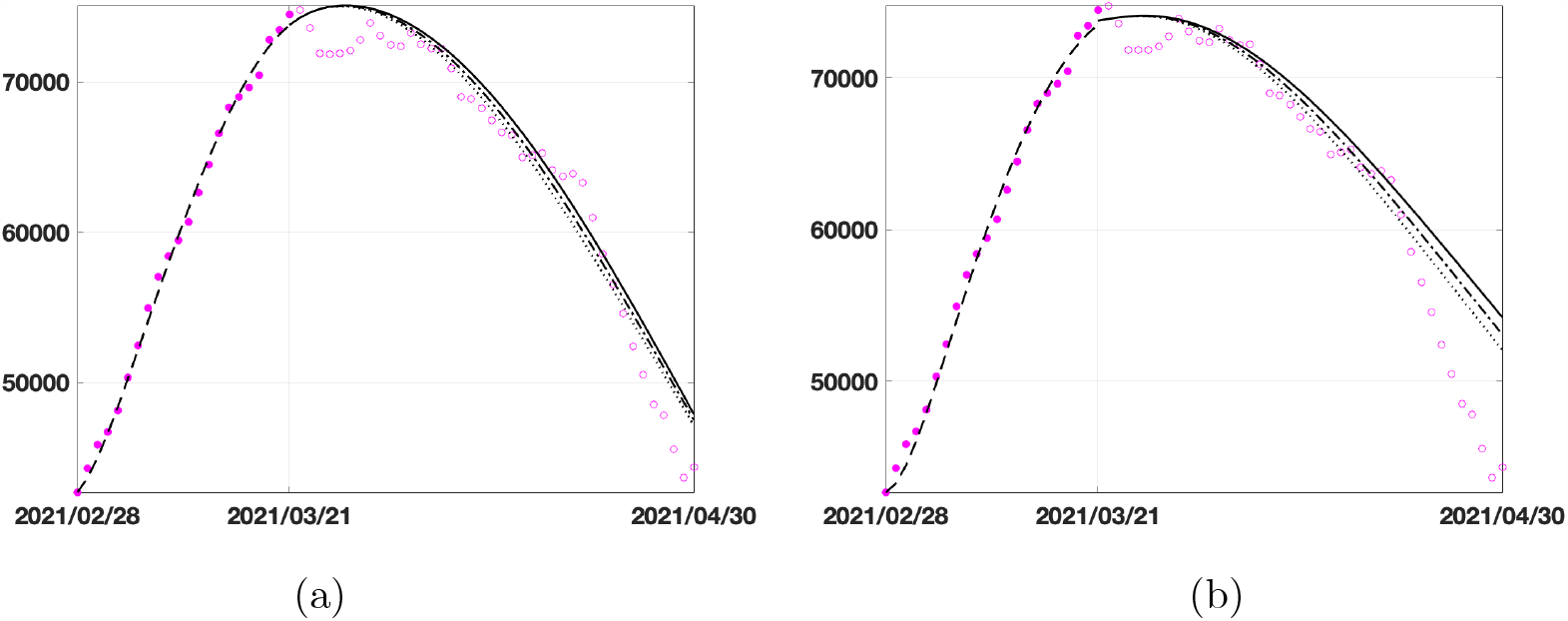
30 days prediction for the Infected compartment in Emilia-Romagna by considering different vaccination rates *v* = 0.0022129 (dark continuous line), *v* = 0.0055323 (dark dashed-dotted line) and *v* = 0.0088517 (dark dotted line) : (a) SEIRDV_ pwl, (b) SEIRDV_pwe

### 3.5. *Towards the* 80% *vaccination*

In this experiment we extend the calibration period to the present date (2021/06/12) (using *L* = 21 as in the previous experiments and *p* = 8) and run the simulations to forecast the time at which 70% − 80% population has received at least the first vaccine dose. As already observed, the two models have a very similar behaviour concerning the fit of vaccinated population and the last estimated vaccination rate is *ν* = 0.0095 for both SEIRDV_pwl and SEIRDV_pwe, equivalent to 584861 administrations per day. Hence we report in figure 13(a) the vaccination data (pink circles), the fitted vaccinated population (black dashed line), together with the forecasts obtained with the calibrated vaccination rate (black continuous line) and with an increased vaccination rate 30% (dot dashed line) and 60% (dotted line) (computed by SEIRDV_pwe). We highlight with dashed lines the values of vaccinated individuals corresponding to 70% and 80% of the whole population. The reproduction number *R*_*t*_ shown in Figure 13(b), obtained by calibrated data, confirms the positive effects of the current vaccination campaign. The progressive decrease of *R*_*t*_ below 1 yields the reduction of the disease spread. The expected similarity between SEIRDV_pwl and SEIRDV_ pwe forecasts is confirmed by data reported in table 4, where the unique small difference appears in the first row. The increase of 60% in the vaccination rate causes a reduction of one month to obtain 70% vaccinated population and about two months (57 *d*) for 80%.

**Table 4:**
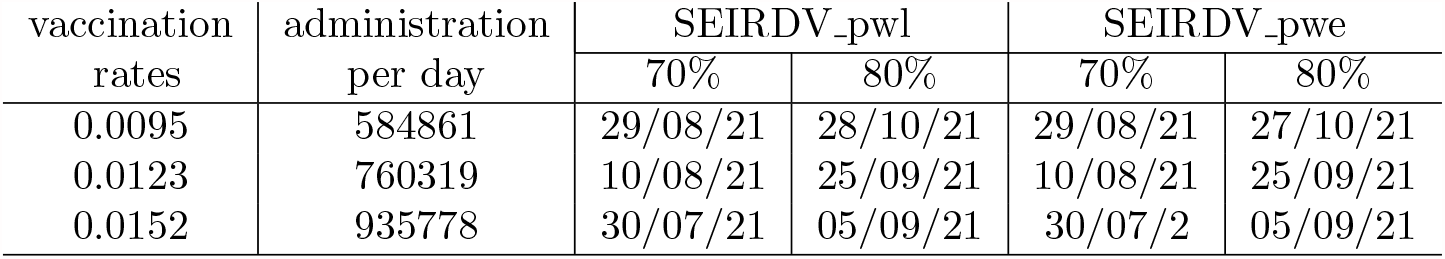
Results of the prediction experiment in Italy obtained by the vaccination rate (first row) calibrated on 12/06/2021 and with a 30% and 60% increase. Dates at which a single vaccine dose is given to 70% − 80% people.

**Figure 13:**
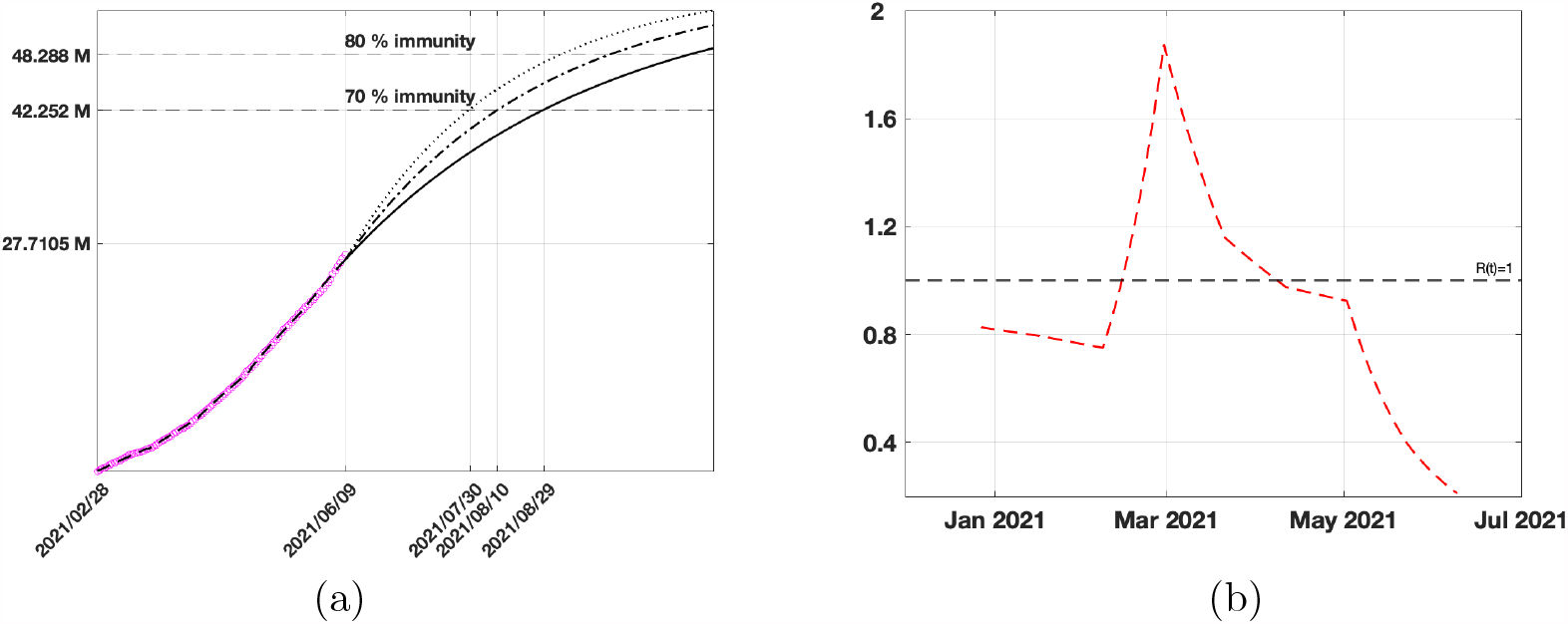
(a) Vaccination data (pink circles), fitted Vaccinated (black dashed line), forecast obtained by SEIRDV_pwe considering the calibrated vaccination rate (black continuous line), an increase of 30% (dot dashed line) and 60% (dotted line) of vaccination rate. (b) Reproduction number *R*_*t*_.

The differences between the two models can be observed in table 5 reporting the number of Infected and Dead individuals in the days of the vaccination targets 70%, 80% reported in table 4. Increasing the vaccination rate *ν*, we observe a reduction of the Dead individuals for each target and both methods. For each target and vaccination rate, the Infected and Dead individuals computed by SEIRDV_pwe are smaller than SEIRDV_pwl.

**Table 5:** Value of Infected and Dead individuals correspondent to 70% and 80% immunization targets for the vaccination rates *ν* displayed in table 4

| Target | $\nu$ | SEIRDV_pwl | | SEIRDV_pwe | |
| --- | --- | --- | --- | --- | --- |
|  |  | I | D | I | D |
| 70% | 0.0095 | 39716 | 129425 | 8890 | 128655 |
|  | 0.0123 | 47873 | 129038 | 18386 | 128552 |
|  | 0.0152 | 55545 | 128785 | 27912 | 128448 |
| 80% | 0.0095 | 15895 | 130048 | 906 | 128739 |
|  | 0.0123 | 19791 | 129621 | 3111 | 128712 |
|  | 0.0152 | 24829 | 129346 | 6713 | 128672 |

The pie graph of Figure 14 shows the percentage of people of all the compartments corresponding to 70% and 80% vaccination targets. In particular Figure 14 (a) represents SEIRDV_pwl reaching 70% vaccination target on 2021/08/29, at the calibrated vaccination rate (*ν* = 0.0095) whereas Figure 14 (b) is relative to SEIRDV_pwe reaching 80% vaccination target on 2021/09/05 at the 60% increased vaccination rate (*ν* = 0.0152). The 10% increase in the Vaccination target causes the same percentage reduction of the susceptible individuals from 23% to 13%, whereas the percentages in the remaining populations do not change significantly. We can consider these pictures as worse and best case predictions of the vaccination campaign in Italy.

**Figure 14:**
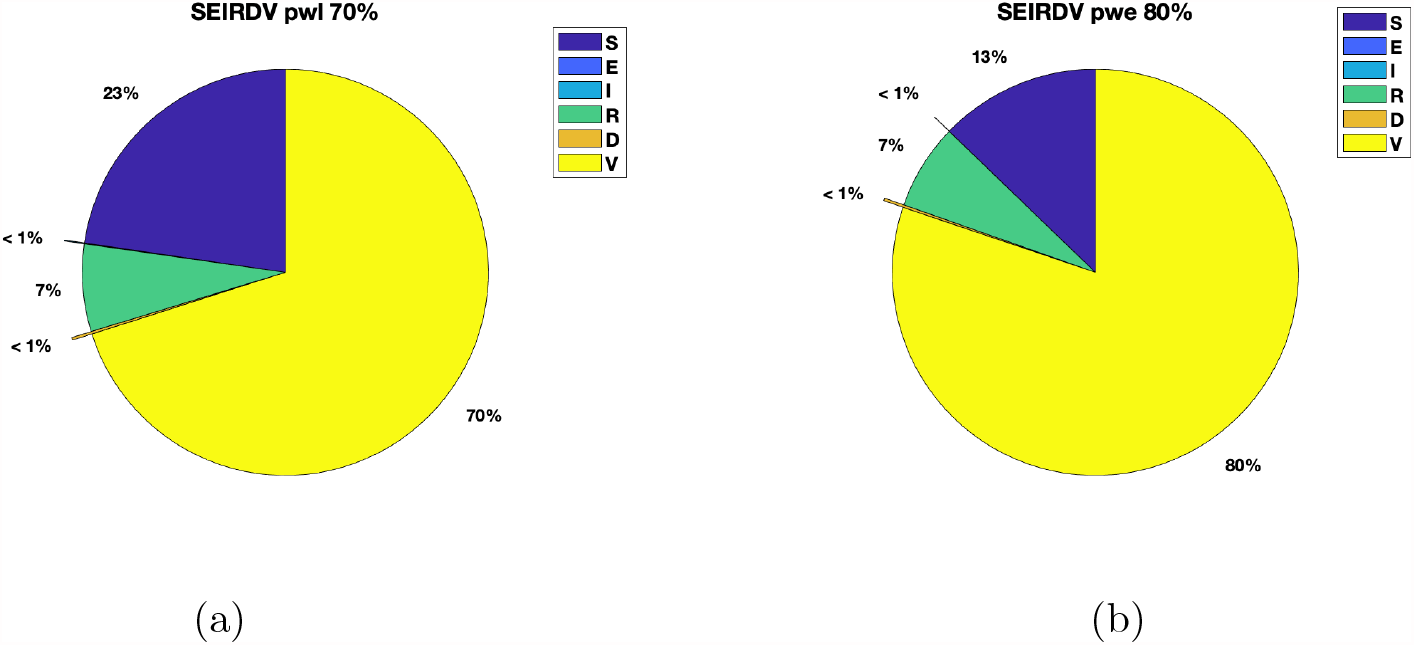
Percentage compartments obtained by SEIRD_ pwl on 2021/08/29 (a) and SEIRD_pwe on 2021/09/05 (b).

## 4. Conclusions

We have proposed two SEIRDV compartmental models, each involving six populations (Susceptibles, Exposed, Infected, Recovered, Dead and Vaccinated) for the analysis of COVID-19 spread during the vaccination campaign in Italy. The two schemes differ in the forcing time-dependent function.

We have calibrated the model parameters in time intervals of about 20 days through a nonlinear constrained least squares minimization. The results on the data on whole Italy and especially on the Emilia-Romagna region are very promising. The data fit obtained is really very faithful with both models for all the considered compartments with a relative residual value less than 1%. The forcing functions, linear and exponential, characterize the model especially when the epidemic spread is increasing.

The simulated predictions of the infected population behaviour in a 30 days period from mid March to mid April 2021 show that the SEIRDV_exp model performs better when compared with the data available in that period. In particular, in the case of Emilia-Romagna we remark that the peak day for the infected population is predicted with great accuracy. Simulations of the Infected curve with a different, increasing, vaccination rate in the same period give the idea of how the epidemic would evolve in these cases.

Finally, we applied the model calibrated up to 2021/06/12 to forecast the epidemic behaviour when 70% − 80% of population has received one vaccine dose. At the present vaccination rate the 80% immunization is reached on 2021/10/28 whereas with an increase of 60% (best scenario) it is reached on 2021/09/05.

Future studies and extensions of the proposed models will consider the limited duration of vaccine-induced immunity and the possible seasonal pattern of the COVID-19 epidemic waves.

## Data Availability

Data are publicly available on the GitHub repository.

https://github.com/fzama63/COVID-SEIRDV

## Appendix A. Vaccines Database Description

We consider the file ‘somministrazioni-vaccini-summary-latest.csv’ in the folder ‘dati’. It contains a table made of 16 fields:

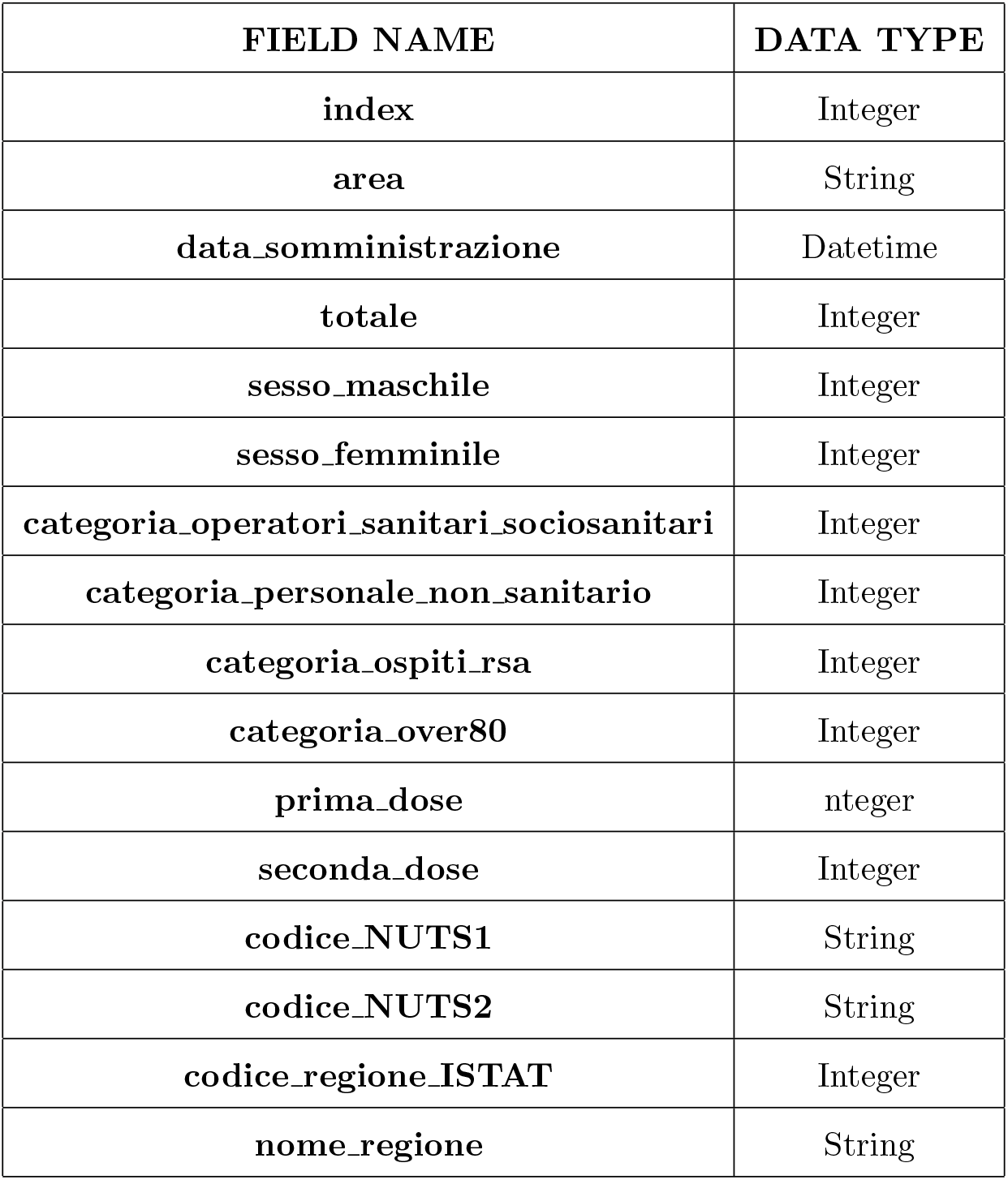

Although some vaccines are administered in two different doses we consider, in our model, as vaccinated the people who received at least the first dose. Since some studies report that the positive immunity effects obtained from a single dose are evident in the immediate days after the vaccine, we can use this information for our short-term prevision.

## Appendix B. Reproduction Number *R*_0_

Concerning the analysis of stability, equilibrium solutions of an SEIR model with different vaccination policies please refer to [22] and references therein. Using the relation *N* = *S*(*t*) + *E*(*t*) + *I*(*t*) + *R*(*t*) + *D*(*t*) + *V* (*t*), we can eliminate the last equation in (1) and define a disease free equilibrium (*S*^∗^, *E*^∗^, *I*^∗^, *R*^∗^, *D*^∗^), with *I*^∗^ = *E*^∗^ = *R*^∗^ = *D*^∗^ = 0. Following the next generation matrix approach [23, 24], we compute the basic Reproduction Number *R*_0_, defined as the number of secondary cases generated by a single Infected. Let *X* = [*E, I*]^*T*^ be the state at infection of system (1), then the Exposed and Infected equations can be written as: *X*′ = *F* (*X*) +*W* (*X*) where

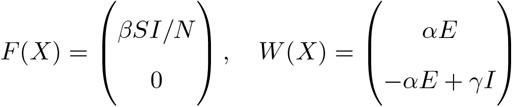

The Jacobian matrices of *F* and *W* at the disease free equilibrium are:

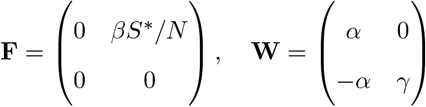

According to [23] the basic reproduction number *R*_0_ is the maximum eigenvalue of the next generation matrix *NGM* = **FW**^−1^, i.e.

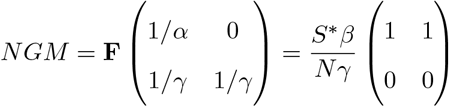

In the assumption that at disease free state *S*^∗^ = *N* we obtain (2). We note that it coincides with the *R*_0_ value of a standard SEIR model [19].

## References

[1] Qingyu Chen, Alexis Allot, and Zhiyong Lu. LitCovid: an open database of COVID-19 literature. Nucleic Acids Research, 49(D1):D1534–D1540, 11 2020.

[2] Kermack W.O. and McKendrick A.G. A contribution to the mathematical theory of epidemics. In Proceedings of the Royal Society of London, volume A, pages 700–721, 1927.

[3] Nicola Parolini, Luca Dede’, Paola Antonietti, Giovanni Ardenghi, Andrea Manzoni, Edie Miglio, Andrea Pugliese, Marco Verani, and Alfio Quarteroni. Suihter: A new mathematical model for covid-19. application to the analysis of the second epidemic outbreak in italy, 2021.

[4] Jie Zhu and Blanca Gallego. Evolution of disease transmission during the covid-19 pandemic: patterns and determinants. Scientific Reports, 11(1):1–9, 2021.

[5] Hamdi Friji, Raby Hamadi, Hakim Ghazzai, Hichem Besbes, and Yehia Massoud. A generalized mechanistic model for assessing and forecasting the spread of the covid-19 pandemic. IEEE Access, 9:13266–13285, 2021.

[6] Giulia Giordano, Marta Colaneri, Alessandro Di Filippo, Franco Blanchini, Paolo Bolzern, Giuseppe De Nicolao, Paolo Sacchi, Patrizio Colaneri, and Raffaele Bruno. Modeling vaccination rollouts, sars-cov-2 variants and the requirement for non-pharmaceutical interventions in italy. Nature Medicine, pages 1–6, 2021.

[7] Satyaki Roy, Ronojoy Dutta, and Preetam Ghosh. Optimal time-varying vaccine allocation amid pandemics with uncertain immunity ratios. IEEE Access, 9:15110–15121, 2021.

[8] Manuel Adrian Acuña-Zegarra Saúl, Díaz-Infante, David Baca-Carrasco, and Daniel Olmos Liceaga. Covid-19 optimal vaccination policies: a modeling study on efficacy, natural and vaccine-induced immunity responses. Mathematical Biosciences, page 108614, 2021.

[9] Nikolaos P Rachaniotis, Thomas K Dasaklis, Filippos Fotopoulos, and Platon Tinios. A two-phase stochastic dynamic model for covid-19 mid-term policy recommendations in greece: a pathway towards mass vaccination. International journal of environmental research and public health, 18(5):2497, 2021.

[10] Rafal Goebel, Ricardo G Sanfelice, and Andrew R Teel. Hybrid dynamical systems. Princeton University Press, 2012.

[11] Liu X. and Stechlinski P. Infectious disease models with time-varying parameters and general nonlinear incidence rate. Applied Mathematical Modelling, 36(5):1974–1994, 2012.

[12] Amine El Koufi, Abdelkrim Bennar, and Noura Yousfi. Dynamics behaviors of a hybrid switching epidemic model with levy noise. Appl. Math, 15(2):131–142, 2021.

[13] Maher Ala’raj, Munir Majdalawieh, and Nishara Nizamuddin. Modeling and forecasting of covid-19 using a hybrid dynamic model based on seird with arima corrections. Infectious Disease Modelling, 6:98–111, 2021.

[14] Elena Loli Piccolomini and Fabiana Zama. Monitoring italian covid-19 spread by a forced seird model. PLOS ONE, 15(8):1–17, 08 2020.

[15] Lauer S.A., Grantz K.H., Bi Q., Jones F.K., Zheng Q., Meredith H.R., Azman A.S., Reich N.G., and Lessler J. The incubation period of coronavirus disease 2019 (covid-19) from publicly reported confirmed cases: Estimation and application. Ann Intern Med., 172(9):577–582, 2020.

[16] Muluneh Alene, Leltework Yismaw, Moges Agazhe Assemie, Daniel Bekele Ketema, Wodaje Gietaneh, and Tilahun Yemanu Birhan. Serial interval and incubation period of covid-19: a systematic review and meta-analysis. BMC Infectious Diseases, 21(1):1–9, 2021.

[17] Fei Zhou, Ting Yu, Ronghui Du, Guohui Fan, Ying Liu, Zhibo Liu, Jie Xiang, Yeming Wang, Bin Song, Xiaoying Gu, Lulu Guan, Yuan Wei, Hui Li, Xudong Wu, Jiuyang Xu, Shengjin Tu, Yi Zhang, Hua Chen, and Bin Cao. Clinical course and risk factors for mortality of adult inpatients with covid-19 in wuhan, china: a retrospective cohort study. Lancet, 2020.

[18] Nithya C Achaiah, Sindhu B Subbarajasetty, and Rajesh M Shetty. R0 and re of covid-19: Can we predict when the pandemic outbreak will be contained? Indian journal of critical care medicine: peer-reviewed, official publication of Indian Society of Critical Care Medicine, 24(11):1125, 2020.

[19] Matt J Keeling and Pejman Rohani. Modeling infectious diseases in humans and animals. Princeton university press, 2011.

[20] Micaela Morettini, Agnese Sbrollini, Ilaria Marcantoni, and Laura Burattini. Covid-19 in italy: Dataset of the italian civil protection department. Data in brief, 30:105526, 2020.

[21] Kenneth P Burnham and David R Anderson. Multimodel inference: understanding aic and bic in model selection. Sociological methods & research, 33(2):261–304, 2004.

[22] Malen Etxeberria-Etxaniz, Santiago Alonso-Quesada, and Manuel De la Sen. On an seir epidemic model with vaccination of newborns and periodic impulsive vaccination with eventual on-line adapted vaccination strategies to the varying levels of the susceptible subpopulation. Applied Sciences, 10(22):8296, 2020.

[23] P. van den Driessche and James Watmough. Reproduction numbers and sub-threshold endemic equilibria for compartmental models of disease transmission. Mathematical Biosciences, 180(1):29–48, 2002.

[24] Fred Brauer, Carlos Castillo-Chavez, and Carlos Castillo-Chavez. Mathematical models in population biology and epidemiology, volume 2. Springer, 2012.

